# LLM-Assisted Taxonomy and Temporal Analysis of Provider Questions About HIV in provider-to-provider telehealth

**DOI:** 10.64898/2025.12.19.25342694

**Authors:** Amir Erfan Zareei Shams Abad, Dima Dandachi, Mirna Becevic

**Affiliations:** University of Missouri Institute for Data Science and Informatics (MUIDSI); University of Missouri, School of Medicine, Missouri Telehealth Network; University of Missouri, School of Medicine, Department of Medicine; University of Missouri, School of Medicine, Department of Dermatology

**Keywords:** HIV, Project ECHO, Telemedicine, Telehealth, Large Language Model

## Abstract

**Introduction:** Ongoing education in HIV care is limited for many healthcare providers working in rural and non-academic settings, which can reduce patients’ access to high-quality care. To guide targeted tele-mentoring and continuing education, we analyzed questions submitted by clinicians during Extension for Community Healthcare Outcomes (ECHO) sessions to characterize learning needs and thematic trends.

**Methods:** We reviewed 78 clinical questions submitted during Project ECHO sessions and developed a structured classification of topics raised by clinicians. Using text-embedding representations and large language models (LLMs), we explored automated approaches to categorize questions and identify thematic clusters. Analyses compared the distribution of topics across professional roles to detect role-specific learning needs.

**Results:** Distinct topic patterns emerged by clinician type. Physicians and pharmacists most often asked about initiating and modifying antiretroviral therapy (ART). Nurse practitioners focused on ART and adherence support, while allied health professionals and PAs raised social-support and care-navigation issues. Medication-related questions frequently highlighted adherence concerns and ART change considerations.

**Discussion:** ECHO questions reveal clear, role-dependent learning needs that can inform targeted tele-mentoring. LLM-based embeddings provided a practical, scalable way to classify questions and monitor trends, supporting more tailored HIV training for different provider groups.

## 1. Introduction

Access to specialized medical care is a major challenge for patients in rural areas, affecting care of patients with complex conditions, such as Human Immunodeficiency Virus (HIV).[1] Advances in antiretroviral therapy (ART) have turned HIV from a fatal disease into a manageable condition.[2] Despite these advances, many people with HIV still face significant barriers to optimal care, often fueled by access challenges.[3] In many parts of the United States (US), especially rural regions, patients struggle with long distances to clinics, transportation issues, and concerns about confidentiality and stigma.[1] A nationwide shortage of clinicians specializing in HIV, expected to worsen as many approach retirement, compounds these issues.[4] This shortage restricts access to high-quality care and strains the healthcare system’s ability to meet the complex needs of a growing, aging population living with HIV.[2][5]

Extension for Community Healthcare Outcomes (ECHO) is a telehealth platform shown to improve capacity for community primary care clinicians to care for patients with costly and complex conditions such as HIV.[6] This case-based tele mentoring approach helps build the skills of local clinicians, improves the quality of care, and supports greater professional satisfaction.[7][8] The effectiveness of the ECHO has been demonstrated across various medical disciplines and geographic regions.[8] The ECHO program has been successful in educating and assisting community physicians in the context of perinatal HIV, leading to better case management and positive patient outcomes, including complete prevention of mother-to-child transmission in a cohort under investigation.[1] Globally, programs like the HIV/TB ECHO in Zambia have enhanced the knowledge and clinical practices of healthcare workers, thereby strengthening local service delivery capacity.[2]

The University of Missouri School of Medicine-based Missouri’s Telehealth Network operates the Show-Me ECHO program to support healthcare providers (HCPs) in managing complex HIV cases. At the core of the program, primary care clinicians submit de-identified cases and specific questions on pre-session forms to HIV experts-hub team. These questions then guide the live-interactive tele-mentoring discussions, ensuring that sessions address the most relevant clinical issues. Analyzing these questions is important because they reveal practical uncertainties and learning needs that attendance counts and surveys often miss. Many HCPs lack adequate continuing education in HIV care, which limits their ability to manage patients and reduces access to specialty-informed treatment in community settings.

To give the ECHO hub an evidence-based perspective on what HCPs ask and when, we created a validated taxonomy of question categories and tracked temporal patterns by profession. Examining themes (such as diagnosis, ART management, comorbidity care, and psychosocial concerns) helps the hub set session priorities and design targeted continuing education. Monitoring how question patterns evolve over time supports program evaluation by indicating shifts in provider confidence or clinical practice. In rural and resource-limited areas, using questions to focus training makes the best use of limited specialist expertise and provides a practical guide for curriculum prioritization, ultimately supporting improved participants’ competency and stronger HIV care in community settings.

## 2. Methods

Since its November 2018 launch, HIV ECHO engaged 424 unique learners (avg. 21 per session), delivered 1,815 instructional hours, and drew participants from 45 Missouri counties (39% of counties) and 12 states; of 414 with repeat-data, 194 (47%) attended two or more sessions. 78 case presentations from 39 providers were retrospectively reviewed and entered <u>into</u> a structured dataset. Patient demographics were recorded, and all pre-session forms and session recordings were abstracted.

Questions were converted to 384-dimensional sentence embeddings using the sentence transformer ‘all-MiniLM-L6-v2’.[9] Pairwise cosine distances between embeddings were used as input to hierarchical agglomerative clustering (HAC). We selected the number of clusters by visually inspecting the dendrogram for natural breaks and by assessing within cluster cohesion and between cluster separation, with clinical interpretability guiding the choice. Given the small sample size (78 cases), finetuning a supervised classifier was not feasible, so we used unsupervised clustering to define the thematic groups.

Each question was initially assigned to a single cluster based on the HAC output. The items in each cluster were then read and reviewed by the study team, and cluster definitions were drafted from the cluster’s contents. This process produced seven clusters (Table 1).

**Table 1.**
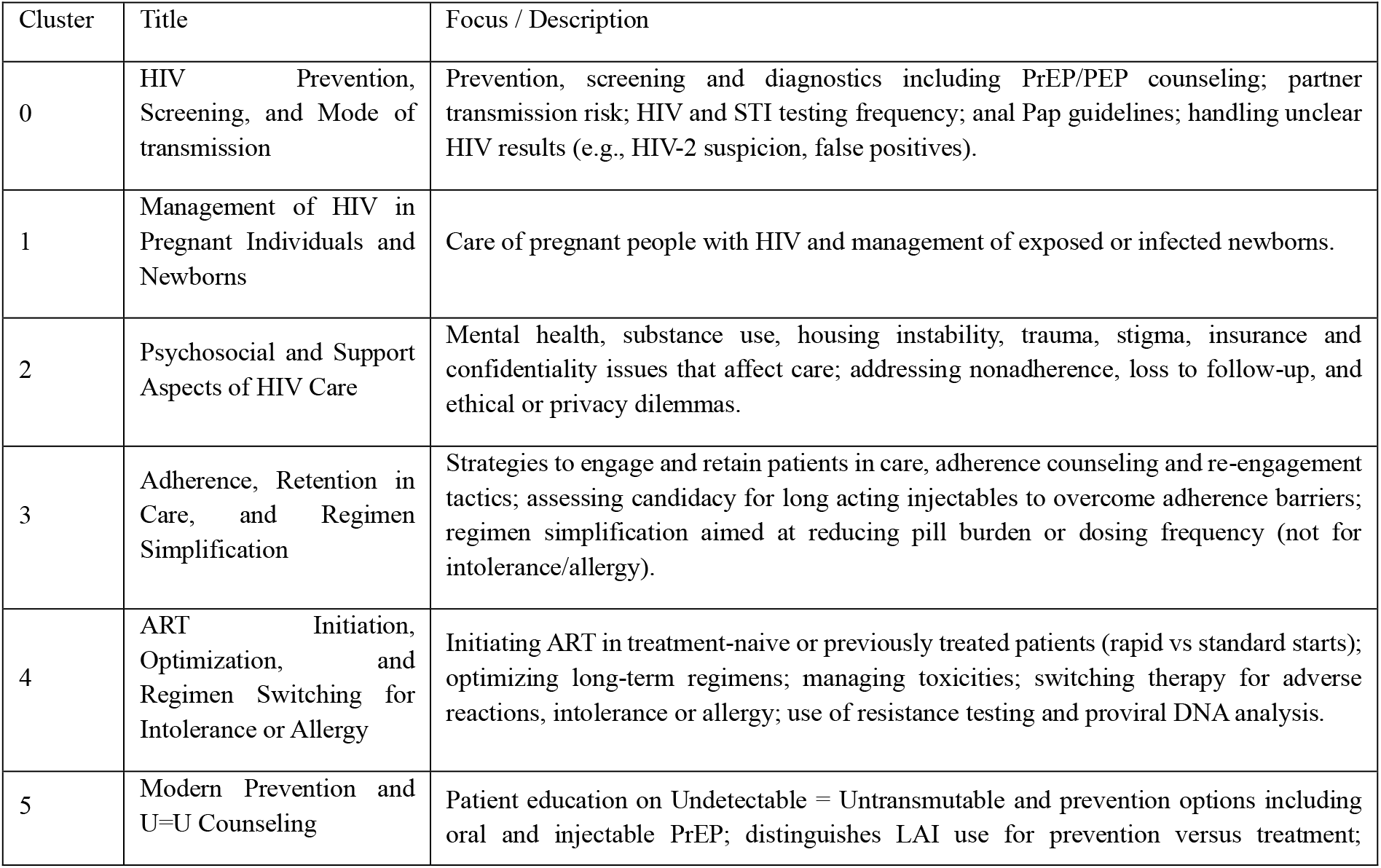
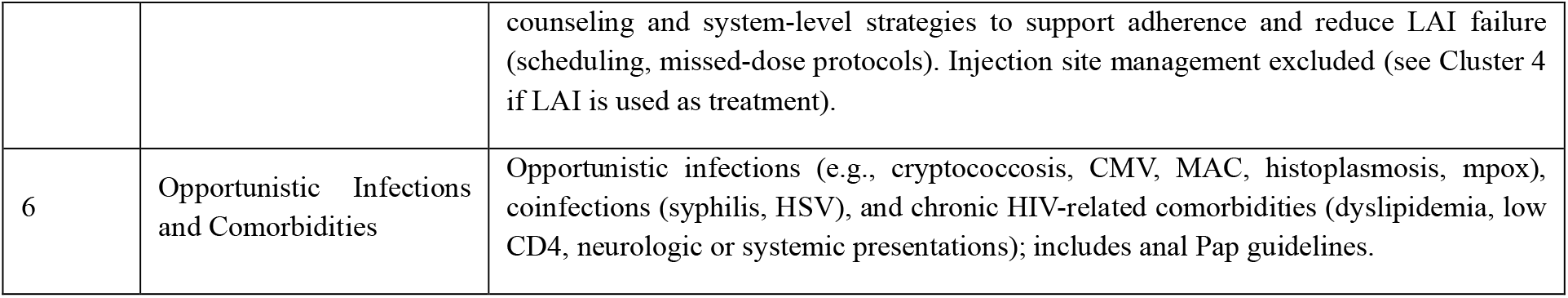
Definitions of seven thematic clusters identified from 78 case questions. Clusters were derived using sentence embeddings from all-MiniLM-L6-v2 and hierarchical agglomerative clustering, and labels were finalized through investigator and pathologist multilabel review.

| Cluster | Title | Focus / Description |
| --- | --- | --- |
| 0 | HIV Prevention, Screening, and Mode of transmission | Prevention, screening and diagnostics including PrEP/PEP counseling; partner transmission risk; HIV and STI testing frequency; anal Pap guidelines; handling unclear HIV results (e.g., HIV-2 suspicion, false positives). |
| 1 | Management of HIV in Pregnant Individuals and Newborns | Care of pregnant people with HIV and management of exposed or infected newborns. |
| 2 | Psychosocial and Support Aspects of HIV Care | Mental health, substance use, housing instability, trauma, stigma, insurance and confidentiality issues that affect care; addressing nonadherence, loss to follow-up, and ethical or privacy dilemmas. |
| 3 | Adherence, Retention in Care, and Regimen Simplification | Strategies to engage and retain patients in care, adherence counseling and re-engagement tactics; assessing candidacy for long acting injectables to overcome adherence barriers; regimen simplification aimed at reducing pill burden or dosing frequency (not for intolerance/allergy). |
| 4 | ART Initiation, Optimization, and Regimen Switching for Intolerance or Allergy | Initiating ART in treatment-naïve or previously treated patients (rapid vs standard starts); optimizing long-term regimens; managing toxicities; switching therapy for adverse reactions, intolerance or allergy; use of resistance testing and proviral DNA analysis. |
| 5 | Modern Prevention and U=U Counseling | Patient education on Undetectable = Untransmutable and prevention options including oral and injectable PrEP; distinguishes LAI use for prevention versus treatment; |
|  |  | counseling and system-level strategies to support adherence and reduce LAI failure (scheduling, missed-dose protocols). Injection site management excluded (see Cluster 4 if LAI is used as treatment). |
| 6 | Opportunistic Infections and Comorbidities | Opportunistic infections (e.g., cryptococcosis, CMV, MAC, histoplasmosis, mpox), coinfections (syphilis, HSV), and chronic HIV-related comorbidities (dyslipidemia, low CD4, neurologic or systemic presentations); includes anal Pap guidelines. |

**Table 2.** Comparative Performance of Large Language Models.

| Model | Precision | Recall | F1-Score | Accuracy (Jaccard) |
| --- | --- | --- | --- | --- |
| Gemini 2.5 Pro (UI) | 0.86 | 0.8 | 0.83 | 0.75 |
| DeepSeek-V3 (UI) | 0.81 | 0.78 | 0.79 | 0.72 |
| GPT-5 Thinking mini (UI) | 0.87 | 0.83 | 0.85 | 0.79 |
| GPT-5 Thinking mini (API) | 0.82 | 0.86 | 0.84 | 0.76 |
| Google Medgemma 27b-text-it | 0.70 | 0.87 | 0.77 | 0.66 |

After cluster definitions were finalized, the questions were manually reviewed and multilabeled by two co-authors, EZ and DD. Each question could receive one or more of the seven cluster labels to reflect multifocal content. These multilabel annotations were used in analyses that examine shifts in focus over time, and by provider role.

To explore the potential of modern LLMs and embedding models for automatic categorization of ECHO session questions, we applied them to the dataset. The labels generated by the models were used only for reporting and comparison purposes and were not incorporated into the study’s main analyses.

Given the limited number of questions, fine-tuning large language models were not feasible. Instead, we applied the finalized cluster definitions in a zero-shot labeling exercise using Four models: Google’s Gemini 2.5 Pro, DeepSeek-V3, GPT-5 Thinking Mini, and Google MedGemma 27B-text-it.[10][11][12][13] Google MedGemma 27B-text-it was selected for its optimization on medical text, which speeds development of healthcare AI applications. Also, Three large language flagship models Google’s Gemini 2.5 Pro, DeepSeek-V3, and GPT-5 Thinking mini — were included in this study because they deliver state-of-the-art performance across many tasks, strong reasoning and generalization, and mature tooling and deployment options, which together provided a practical balance for unsupervised, zero-shot labeling.

The group definitions from Table 1 were supplied to each model. Automated multilabel inference was performed using the chat interfaces of Google’s Gemini 2.5 Pro, DeepSeek-V3, and GPT-5 Thinking mini. Google/MedGemma 27B-text-it — which does not expose an API — was loaded from Hugging Face and executed in Google Colab on an NVIDIA A100 GPU.

Although different outputs can sometimes be produced by application programming interface (API) calls and chat-based user interfaces (UIs) (due to hidden system prompts, sampling defaults, or UI post-processing), the labeling results were found to be very similar. This similarity is attributable to the use of the same underlying model weights where available, the provision of short, prescriptive prompts, and inference being run with effectively deterministic settings (e.g., low temperature and structured JSON output). For comparing UI responses and API responses, the GPT-5 Thinking Mini API was also used, and the results from that model’s API were examined alongside ChatGPT UI outputs. Since the temperature setting for the ChatGPT UI is not publicly known, the API temperature was set to 0.7 to introduce a comparable level of creativity and better mimic UI-like responses. UI post-processing was minimized, and identical post-processing rules were applied to every output. Beyond thematic categorization, patterns of adherence were also examined, as adherence remains a central issue in HIV care. Information was drawn from pre-session forms completed by primary care clinicians, which outlined patient cases, and the Echo session recordings, where clinicians presented their questions to specialists. Each case was assigned an adherence status of good, poor, missing, or N/A. It’s important to note that the “N/A” label was most frequently applied to individuals who had just started treatment or who had not yet been diagnosed with HIV. It does not indicate missing data.

## 3. Results

A total of 78 case presentations were included in the analysis. Patient demographics showed a predominance of males (77%) with females making up the remaining 23%. The age distribution of patients is illustrated in Figure 1 a. In addition to patient characteristics, the dataset includes the professional credentials of the HCPs who submitted questions during the sessions. To facilitate role-based analysis of participation and case presentations, attendees were grouped into four professional categories based on their credentials: Physicians, Nurse Practitioners (NPs), Pharmacists (PharmD), and All other credentials were grouped as Other Allied Health professionals. Physicians included both Medical Doctors (MDs) and Doctors of Osteopathic Medicine (DOs), Nurse Practitioners encompassed credentials such as FNP-C, NP-C, FNP, APRN, and DNP, while Pharmacists included those with PharmD or Pharm.D. degrees.

**Figure 1.**
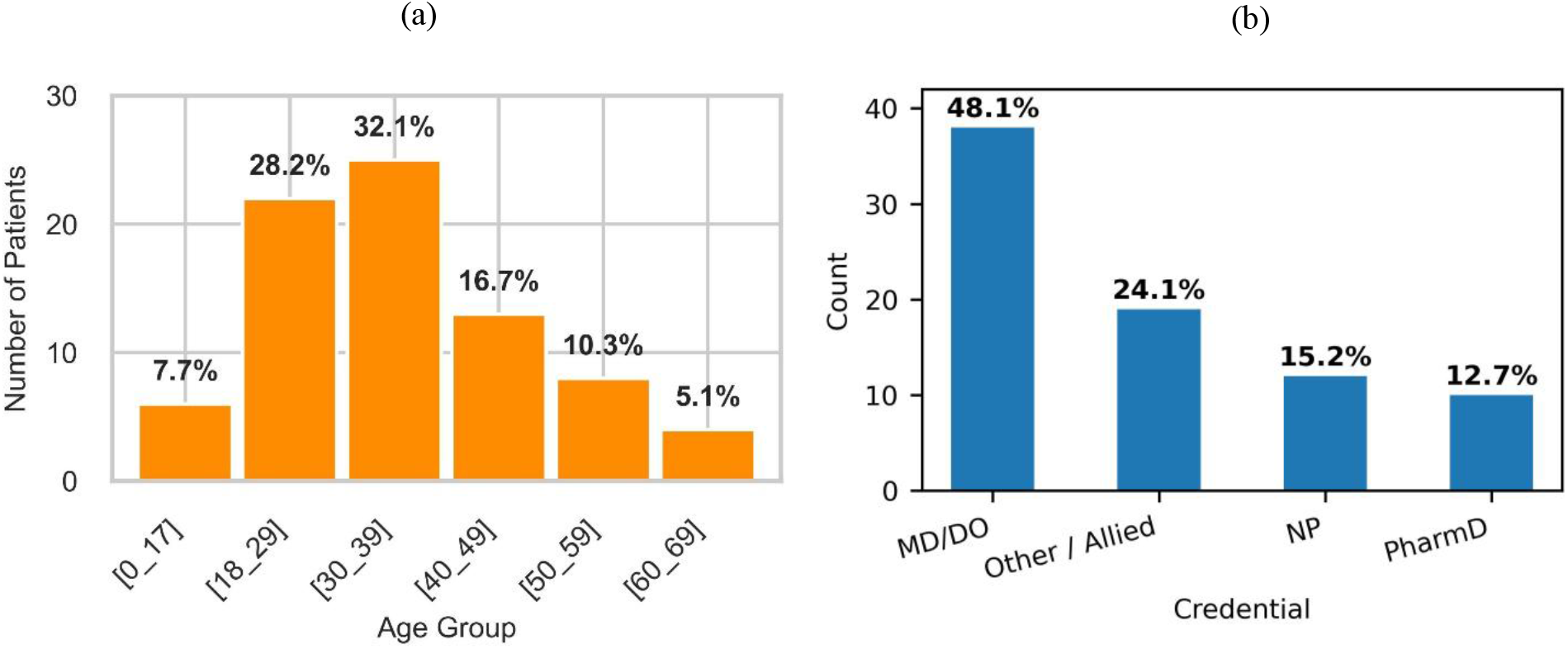
Age distribution of patients represented in case presentations from 2018 to 2025 on Figure 1 (a). Figure 1 (b) is illustrating Distribution of professional credentials among primary care clinicians who submitted questions during ECHO sessions.

**Figure 2.**
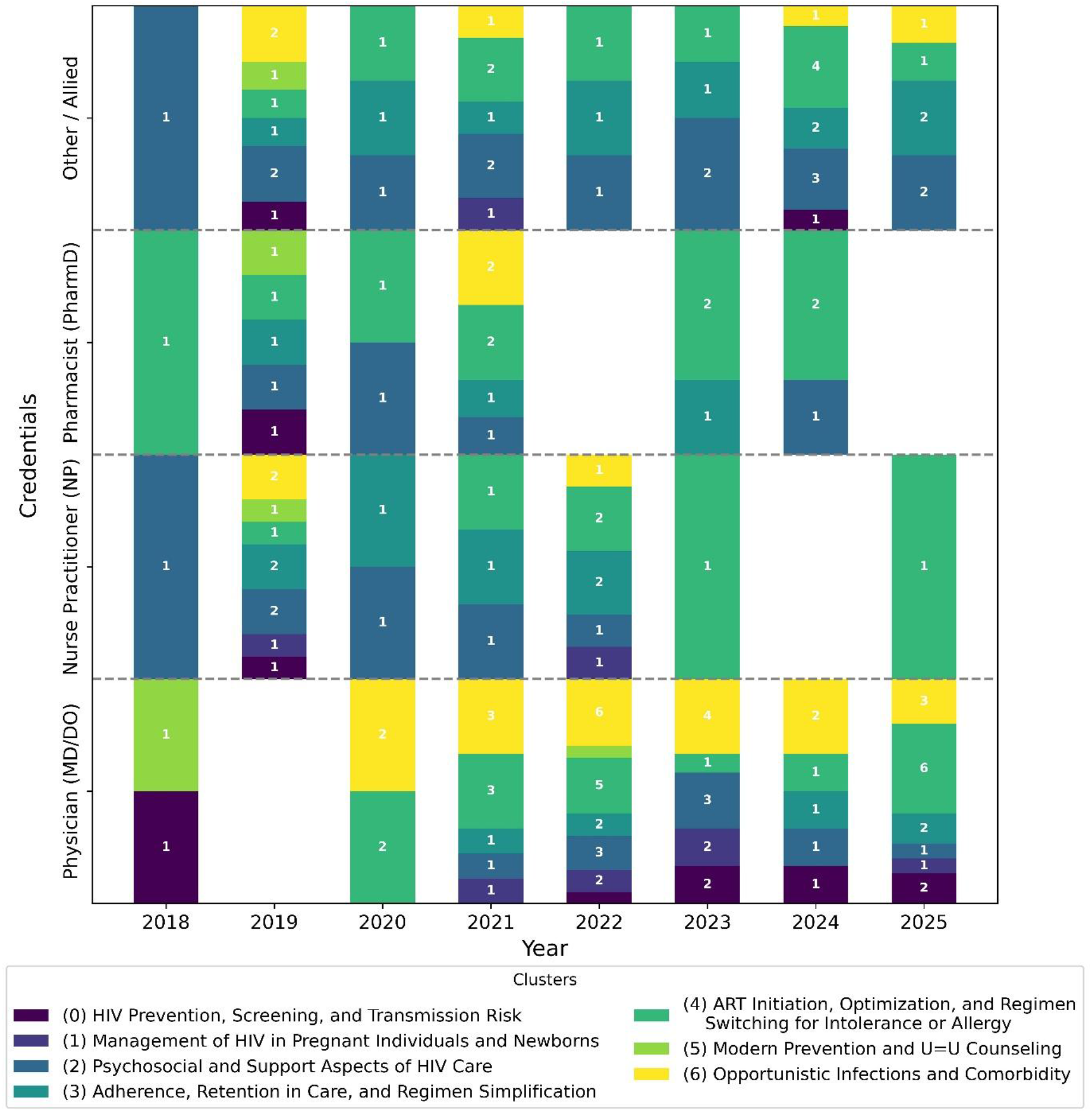
Temporal trends in the thematic composition of questions asked by healthcare providers from 2018 to 2025

These groupings were selected to reflect differences in clinical authority and practice independence. In Missouri, both MD/DO physicians and Nurse Practitioners can prescribe medications, though Nurse Practitioners typically do so under collaborative practice agreements, whereas MDs and DOs practice independently. Pharmacists and Physician Assistants, while integral to patient management and ECHO participation, have different scopes of practice and generally cannot prescribe independently. Distinguishing these roles allowed for a more accurate understanding of the professional composition of the HIV ECHO community and the relative contributions of each discipline to case-based learning and knowledge sharing. The distribution of these provider credentials is shown in Figure 1.b, offering context for the mix of professional backgrounds represented in the ECHO program. Additional details on session history, including provider participation and case counts, are provided in Appendix A.

Overall engagement increased over time, with physicians and allied professionals driving most of the rise. The subject-matter composition shifted over the study period: early activity was dominated by psychosocial/support topics, while later years show growing emphasis on ART initiation/optimization and on opportunistic infections/comorbidity care. Figure 3 presents the topic clusters alongside provider groups, enabling comparison of the types of questions asked in ECHO sessions during the study period.

**Figure 3.**
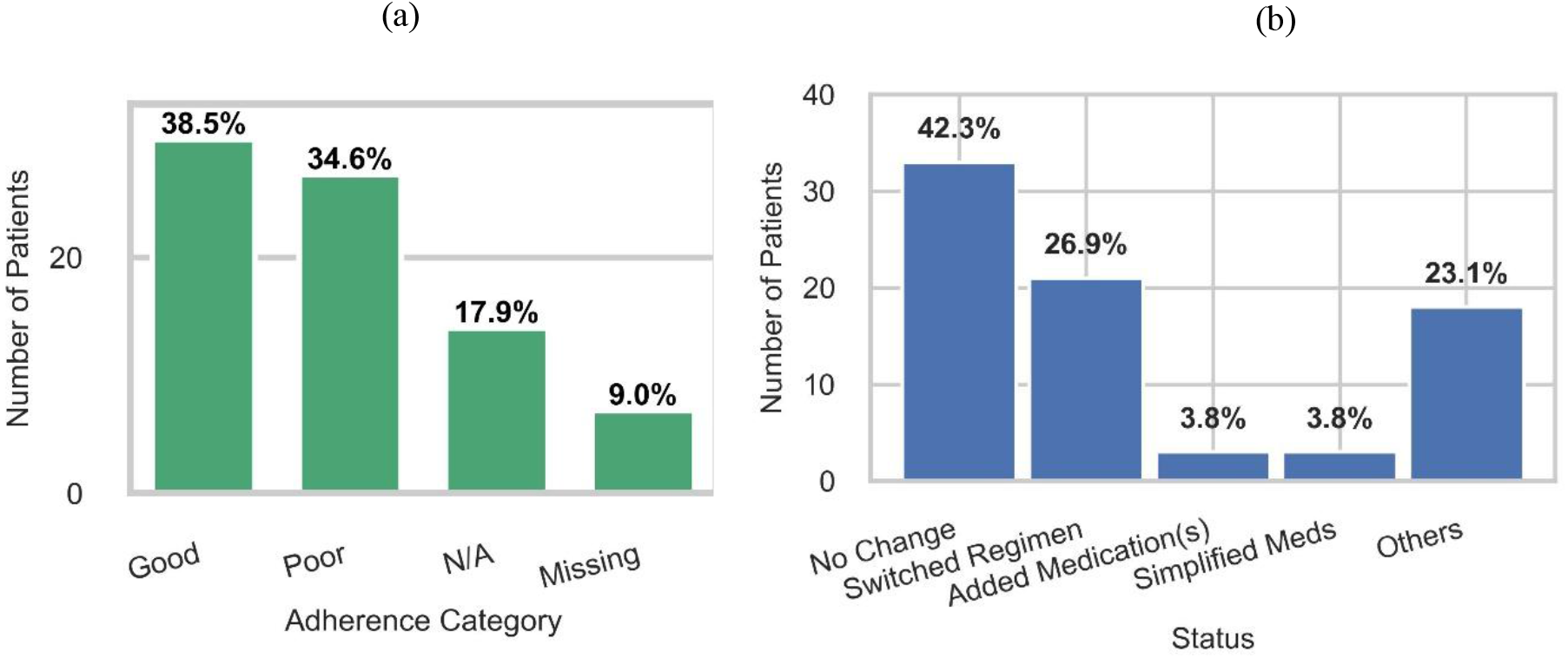
Figure 3a. Distribution of patient adherence statuses in the ECHO program, including good, poor, missing, and N/A categories. Figure 3b. Patterns of antiretroviral medication changes among patients, showing the proportion who switched, did not change, had medications added, had regimens simplified, or fell into other categories (e.g., Negative HIV, Not Started, Not Available, New Start, or Data Missing).

Throughout the study period, Cluster 4, ART initiation/optimization, was the main topic of for physicians. There was a noticeable increase in Cluster 6 questions in subsequent years, reflecting growing attention to opportunistic infections and comorbidities. In the most active years, questions centered on ART and OI predominated, with sporadic appearances of psychosocial/support barriers (Cluster 2) and adherence/retention tactics (Cluster 3).

Nurse practitioners showed an early peak in Cluster 2 (psychosocial/support barriers) around 2018. In the middle years, their activity was split between prevention/screening and adherence topics. Notably, between 2023 and 2025, although the contribution of NPs significantly decreased, all questions fell within Cluster 4 (ART initiation and optimization). This indicates that while concerns regarding adherence and opportunistic infections persisted, there was an increasing emphasis on treatment initiation and regimen management.

Although pharmacist involvement was inconsistent, it mostly focused on Cluster 4 (ART initiation/optimization), which was prominent in 2018 and in the following years like 2020 and 2024. Pharmacists also show some activity in Cluster 2 (psychosocial/support) and Cluster 3 (adherence/retention), but ART regimen issues dominate their contributions.

Other allied professionals consistently emphasized Cluster 2, psychosocial and support barriers, throughout the study period. This emphasis was strong in 2018 and remained elevated in the years that followed. There is a gradual increase in clinical topic clusters (Cluster 3 and 4) toward the later years, but psychosocial and structural support topics remain in their primary focus.

The ECHO program provides information on patient adherence, including patterns of medication use, treatment history, and current regimens. In our sample, 38.5% of cases were reported to have good adherence, while 34.6% of cases were reported to have poor adherence. The remaining cases were labeled as missing or N/A, where “N/A” refers to patients who had recently started treatment or had not yet received an HIV diagnosis. These patterns are summarized in Figure 3.a, which illustrates the full distribution of adherence statuses across the 78 cases.

Patterns of medication changes were also examined. Among the cases, 26.9% of patients had completely switched their antiretroviral regimen at some point during their treatment, 42.3% had no change to their medication, 3.8% had medications added to their existing regimen as recommended by the hub team, and 3.8% had their medications simplified by the hub team. Patients who did not fit these categories were grouped as Other, including those labeled as Negative HIV, Not Available, New Start/No Prior Data, or missing data 3.b).

To explore the potential of LLMs for automatically categorizing ECHO session questions, we evaluated zero-shot classification performance between model generated labels and the manually assigned multilabel clusters. 4 models were compared: Google’s Gemini 2.5 Pro, DeepSeek-V3, Google/MedGemma 27B-text-it, and GPT-5 Thinking mini.

Among the evaluated models, GPT-5 Thinking mini (UI) performed best overall, achieving the highest F1-score (85%) and accuracy (79%). Gemini 2.5 Pro (UI) followed closely with an F1-score of 83% and an accuracy of 75%. The API version, GPT-5 Thinking mini (API), also demonstrated strong performance with a high recall of 86% and an F1-score of 84%. DeepSeek-V3 (UI) showed solid performance as well, with an F1-score of 79% and an accuracy of 72%. Although Google Medgemma 27b-text-it attained the highest recall of all models (87%), its lower precision (70%) resulted in an F1-score of 77% and the lowest accuracy (66%).

## 4. Discussion

Many HCPs lack sufficient continuing education in HIV care, which limits their ability to manage patients and reduces access to specialty-informed treatment in community settings. In this study, de-identified ECHO session questions were analyzed using sentence embeddings and hierarchical agglomerative clustering to identify profession-specific trends over time and to create a structured taxonomy of clinician queries. This approach provides insight into the types of questions clinicians ask, the timing of those questions, and how their priorities change over time. Such insights can guide the tailoring of continuing education curriculums and the prioritization of virtual training content. This framework is scalable to other ECHO programs; it offers a practical foundation for aligning curricula with clinician needs and improving the relevance of case-based learning.

The patterns observed across different HCPs groups highlight clear priorities for ECHO sessions. For physicians and pharmacists, the focus should be on regimen selection, treatment initiation, and optimization. For nurse practitioners, greater attention to prevention, U=U counseling, and practical support for adherence is needed. For allied staff, maintaining a strong emphasis on psychosocial issues, structural barriers, and care navigation remains important. Aligning the ECHO curriculum with these areas can make the sessions more relevant and improve their impact across Missouri’s diverse HIV care workforce.

In addition to the thematic trends, this study examined patient level data from the ECHO program. Patterns of antiretroviral medication changes, with nearly one-third of patients switching therapy, while others either remained on their current regimens, added to existing medications, or underwent simplification. With pattern recognition techniques ECHO datasets could be leveraged to identify clinically meaningful subgroups and associations—for example, whether patients represented in the ART initiation/optimization group has distinct patterns in their medication history. This type of analysis could support a more detailed knowledge of HIV care delivery in real world settings.

The potential of LLMs and embedding-based methods for categorizing and analyzing ECHO questions warrant further exploration. Other ECHO programs run on a much larger scale, with thousands of cases available for study. The methods demonstrated here provide a starting framework for such programs: Clustering and zero-shot labeling can provide actionable insights even with small datasets. Large datasets could enable finetuning of LLMs to automatically and accurately multilabel clinician questions. Results from this study suggest promise in that direction, with GPT-5 Thinking Mini achieving 79.0% accuracy. As ECHO datasets expand, machine learning and LLM-based methods could play a growing role in analyzing clinician needs and aligning tele-education with the changing landscape of HIV care.

There are several intriguing trade-offs in the model behavior when comparing the ChatGPT UI and API outputs for GPT-5 Thinking Mini. While the API showed higher recall, suggesting it catches a wider set of pertinent items but includes a few more false positives, the UI showed higher precision, suggesting it tends to give more conservative outputs with fewer false positives. This difference may reflect the fact that the UI has hidden inference settings and potential internal post-processing, whereas the API was run with a set temperature of 0.7 to approximate UI-like creativity. Although these settings add some variability, the labeling patterns are generally consistent, as evidenced by the slightly lower F1-score and accuracy for the API. These findings underline the significance of assessing outputs in the context of the intended application by showing that, even though the underlying model weights remain the same, interface-specific elements as sampling randomness, prompt handling, and post-processing can affect performance.

## Data Availability

All data produced in the present study are available upon reasonable request to the authors

## Acknowledgments

The authors would like to thank the Missouri Telehealth Network Knowledge Management team for their help with HIV ECHO attendance data analysis.

## Author Contributions

Conceptualization, Methodology, Data Curation, Formal Analysis, Investigation, Project administration, Visualization, Software, Writing – Original Draft, and Writing – Review & Editing: Amir Erfan Zareei Shams Abadi

Conceptualization, Investigation, Resources, Supervision, Validation, Writing – Review & Editing: Dima Dandachi

Conceptualization, Investigation, Resources, Supervision, Validation, Writing – Review & Editing: Mirna Becevic

## Appendix A Session History: Provider Participation & Case Counts

**Table A1.** Summary of sessions showing provider attendance and number of cases discussed per session.

| YEAR | Case presentations | percent | Cumulative frequency | Cumulative percent | Sessions (n) |
| --- | --- | --- | --- | --- | --- |
| 2018 | 3 | 3.8% | 3 | 3.8% | 3 |
| 2019 | 10 | 12.8% | 13 | 16.7% | 6 |
| 2020 | 5 | 6.4% | 18 | 23.1% | 6 |
| 2021 | 12 | 15.4% | 30 | 38.5% | 15 |
| 2022 | 15 | 19.2% | 45 | 57.7% | 13 |
| 2023 | 11 | 14.1% | 56 | 71.8% | 14 |
| 2024 | 11 | 14.1% | 67 | 85.9% | 16 |
| 2025 | 11 | 14.1% | 78 | 100.0% | 9 |

